# A Two Arm, Open Label, Randomized, Multi centric, Interventional, Prospective, Clinical Study to Evaluate Efficacy and Safety of Nicotine Free Ayurvedic Formulation (SMOTECT-AZAADI granules) in comparison with Nicotine Replacement Therapy (NRT) in Tobacco users (Chewing)

**DOI:** 10.1101/2025.06.02.25328797

**Authors:** Sanjay Tamoli, Vaishali Deshpande, Shishir Pandey, Narendra Mundhe, Santosh Irayya Swami, Eshwari Salian, Kamalesh Mahajan, Sandip Birari, Shruti Tarapure, Ruby Dubey, Ninad N. Mulye, Gurseet Singh

## Abstract

**Background:** Smokeless tobacco (SLT) poses a major public health risk, linked to cancer, heart disease, and stroke. Nicotine Replacement Therapy (NRT) is commonly used for tobacco cessation. However, it carries a high risk of relapse, which significantly limits its long-term effectiveness. Nicotine free Ayurvedic formulations (NFAF), having potential anti-stress and mood enhancing properties may offer a healthier promising alternative.

**Purpose:** The study evaluated the efficacy and safety of the proposed Nicotine free Ayurvedic formulation compared to NRT in tobacco chewers.

**Methods:** A two-arm, open-label, randomized, multicentric clinical trial involving 112 participants was conducted with one group receiving NFAF and the other NRT for 90 days with follow ups every 15 days till day 90 and a follow up on day 120 after 30 days of non intervention. Parameters such as frequency, duration of tobacco/gutka usage to evaluate dependency on nicotine, mouth opening capacity, quality of life, physiological measures and clinical symptoms were assessed. Safety assessment was also done based on clinical and laboratory parameters.

**Results:** The proposed Nicotine free Ayurvedic formulation was observed to be as effective as NRT in reducing tobacco consumption and dependence. Significant and comparable reduction in stress, fatigue and improvement in energy strength, stamina and well-being were observed with the use of NFAF. Clinical symptoms such as headache, fatigue, and mood changes improved similarly in both groups. A higher percentage of NFAF users (21.56%) achieved complete cessation of tobacco consumption versus NRT users (17.77%). NFAF was observed to be well tolerated, with only minor, self-limiting adverse events.

**Conclusion:** NFAF effectively reduced SLT dependence and improved quality of life and clinical symptoms, with good safety. It represents a healthier alternative for tobacco cessation.

## Introduction

Tobacco is being used all over the world for various purposes etc.^1^ India is the second largest producer as well as consumer of tobacco next only to China^2^ . The term smokeless tobacco (SLT) refers to the consumption of unburned tobacco, in the form of chewing, snuff, etc. Recent national data from the Global Adult Tobacco Survey shows that 21.4% of adults in India are current users of smokeless tobacco.^3^ Among the chemical components identified in SLT, 28 have been observed to be carcinogenic, with nitrosamines being the most significant.^4^ Its regular use has been strongly linked to the development of oral cancer and other oral conditions such as leukoplakia and gum recession. Nicotine absorbed through the oral tissues may cause addiction and physical dependence.^5^

As of 2015, Gutka is regarded as the fourth most common addictive product worldwide. Consumers of gutka believe that it acts as a digestion aid, exhibits antimicrobial properties and provides a sense of well-being.^4^ However, is a known carcinogenic, causing cancers of the mouth, neck, throat, esophagus and other parts associated with the gastrointestinal tract.^6^ Gutka consumption during pregnancy has been found to significantly increase the risk of maternal anemia and is associated with a higher incidence of adverse neonatal outcomes.^7^ Gutka consumption also causes oral and dental disorders.^8^

Cessation of tobacco chewing is the best way to significantly reduce the risk associated with it. However, it may cause withdrawal symptoms such as anxiety, irritability, dysphagia, hedonic deregulation, etc. if done abruptly. Measures such as behavioral treatment, nicotine replacement therapy (NRT) and pharmacological treatment are being used to aid in tobacco cessation presently. Behavioral treatment is the basic line of treatment, which includes consumer education and social support. NRT utilizes products such as nicotine chewing gums, transdermal patches, nasal nicotine solution, and nicotine vapor inhalers. Pharmacological therapies include the use of anti-depressant drugs and other symptomatic management measures. NRT has partial success rate. The pharmacological strategies currently in use alleviate the withdrawal symptoms but also exhibit various side effects. ^9,10^

The proposed nicotine free Ayurvedic formulation (NFAF) is intended to be useful in the cessation of tobacco chewing (e.g., Gutka). To validate this claim, a clinical study was conducted with the objective of evaluating the efficacy and safety of this formulation in comparison with NRT in chewing tobacco users through a two-arm, open-label, randomized, multi-centric, interventional, prospective clinical design.

## Materials and methods

### Study design and sites

This two arm, open label, randomized, multi centric, interventional, prospective, clinical study was conducted at four sites:

1. KVTR Ayurved College and Hospital, Shirpur, Dhule-424001
2. Parul Institute of Ayurveda & Research Post. Ishwarpura-Limba, Waghodia, Vadodara, Gujarat-391760, India
3. Department of Kayachikitsa, Seth Govindji Raoji Ayurved Mahavidyalaya, Solapur-413001
4. Ayurved Seva Sangh Ayurvedic College & Hospital, Ganeshwadi, Panchavati, Nashik, Maharashtra - 422003, India

### Ethical considerations and informed consent

Study was conducted after approval from the Institutional Ethical Committees of the respective study sites-

KVTR Ayurved College and Hospital – AMB/516/2023-24

Parul Institute of Ayurveda & Research – PAIR/IEC/04/2023

Seth Govindji Raoji Ayurved Mahavidyalaya – IEC/LTR/SGR/04/2023-24

Ayurved Seva Sangh Ayurvedic College & Hospital – 646

CTRI registration was done with registration number - CTRI/2023/07/055111 on: 12/07/2023. Consenting participants meeting the inclusion criteria were considered for the study. The study was carried out and reported adhering to CONSORT statement.

### Details of intervention

NFAF granules are an Ayurvedic Proprietary Medicine containing *Kauncha* (*Mucuna pruriens*) seed extract, *Ashwagandha (Withania somnifera)* root extract, *Yashtimadhu* (*Glycyrrhiza glabra*) root extract, *Lavang (Syzygium aromaticum)* flower bud powder, green tea (*Camellia sinensis)* leaf extract, *Akalkara* (*Anacyclus pyrethrum*) root powder, guava (*Psidium guajava)* fruit, *Supari* (*Areca catechu*) seed, *Amaltas (Cassia fistula)*, and *Tulasi* (*Ocimum tenuiflorum*) leaf extract. The product is manufactured by Smotect Pvt. Ltd, Mumbai, India and is available in the market by the name of SMOTECT – AZAADI granules.

### Study dosage, duration and visits

The study involved two groups; one group was administered 5 gm NFAF granules twice daily after food for 90 days. The other group was asked to take NRT dosed 2 mg twice daily for 90 days.

Interventions were administered for a total duration of 90 days with a 30 day follow up period without administration of intervention. The study visits were planned and various outcomes were assessed on Screening visit, baseline visit (day 0) and days 15, 30, 45, 60, 75, 90, 120 (Visit at Day 120 was a follow up telephonic visit to evaluate relapse of use of tobacco).

### Inclusion criteria

Males and females between the ages of 18 and 70 years (inclusive) without any significant medical or surgical illness having a habit of chewing tobacco (e.g., gutka) at least three times daily for a minimum duration of one year were considered. All participants were required to be willing to comply with the study procedures and voluntarily provide written informed consent.

### Exclusion criteria

Subjects with any known severe, chronic or infectious diseases, active malignancy, history of a significant cardiovascular event within 12 weeks prior to randomization; and active metabolic/gastrointestinal diseases that could interfere with nutrient metabolism (except diabetes) were excluded. Individuals with uncontrolled diabetes or hypertension, those who had participated in any clinical study, or those with known hypersensitivity to any component of the study drug were excluded along with pregnant/ lactating females, and individuals who were deemed unsuitable for participation by the investigator for reasons not explicitly listed were also excluded from the study.

### Sample size

A total of 116 participants were screened in the study of which 112 were randomized and there were 04 screen failures. The details of participant enrollment, allocation, follow-up, and analysis are illustrated in the CONSORT flow diagram.

**Figure 1:**
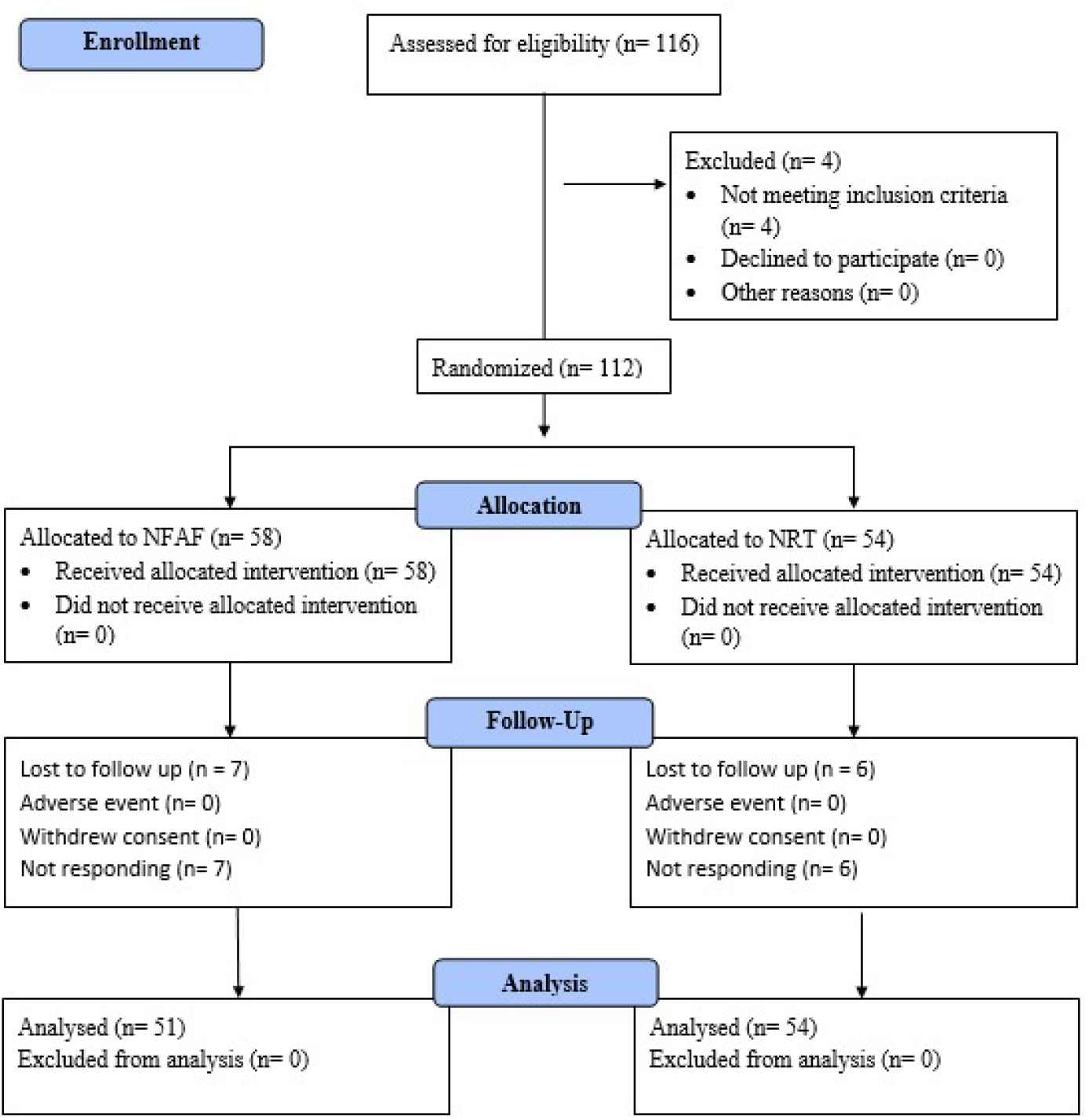
Consort diagram.

### Outcome measures

Primary outcome measures under consideration were the frequency and duration of tobacco consumption. Secondary outcome measures included mouth opening capacity, quality of life (QOL) assessed using WHO QOL BREF^11^, stress levels using Perceived Stress Scale^12^, sleep disturbances using Insomnia Severity Index (ISI)^13^, physiological parameters, clinical symptoms related to tobacco chewing, nicotine dependence using Fagerström Test^14^, relapses of tobacco consumption after stopping the interventions, urine nicotine level and safety related parameters.

### Statistical analysis

Statistical analysis was performed Graphpad statistical software. For the analysis of efficacy variables, data was analyzed from the Intent to treat population and per protocol population also. The values of the last visit were considered for final analysis for participants who did not complete the study schedule (Last Observation Carry Forward) for intent to treat analysis. Safety Analysis was done on all participants who have administered at least one dose of treatment. Data describing quantitative measures were expressed as mean ± standard deviation or standard error or the mean with range. Qualitative variables were presented as counts and percentage. Comparison of variables representing categorical data was performed using t test and Chi-square test. All *P* values were reported based on two-sided significance, and all the statistical tests were interpreted at least up to 5% level of significance.

## Results

### Baseline demography

Both groups were comparable in terms of age, weight, BMI, and tobacco consumption habits.

**Table 1:** Baseline demography.

| Parameter | NFAF group | NRT Group |
| --- | --- | --- |
| **n** | 51 | 48 |
| **Male** | 40 (78.43%) | 41 (85.41%) |
| **Female** | 11 (21.56%) | 7 (14.58%) |
| **Age (Mean SD)** | 37.96 ± 12.92 (19-66) | 38.83 ± 9.92 (19-66) |
| **Weight** | 64.96 ± 15.36 | 67.92 ± 12.23 |
| **BMI** | 24.85 ± 5.35 | 24.83 ± 3.91 |
| **Gutka** | 17 (33.33%) | 16 (31.25%) |
| **Tobacco** | 30 (58.82%) | 31 (64.58%) |
| **Mishri** | 2 (3.92%) | - |
| **Gutka + Tobacco** | 2 (3.92%) | 1 (2.08%) |
| **History of consumption (Mean SD)** | 10.72 ± 9.44 | 9.91 ± 7.68 |
| **Average Daily consumption (Mean SD)** | 6.94 ± 3.65 | 7.23 ± 3.84 |
| **Time of chewing on every consumption (Mean SD)** | 14.08 ± 8.71 | 14.63 ± 9.18 |
| **Hypertension** | 2 | 2 |
| **Diabetes Mellitus** | 0 | 1 |
| **Hyperthyroidism** | 1 | 0 |
| **Hypothyroidism** | 1 | 0 |

### Frequency of tobacco consumption

NFAF group exhibited a significant reduction in daily tobacco chewing frequency, from 7.23 ± 3.84 at baseline to 2.41 ± 2.25 at day 90, which was comparable to the NRT group, where the frequency reduced from 6.94 ± 3.65 to 2.27 ± 2.07 over the same period.

**Graph 1:**
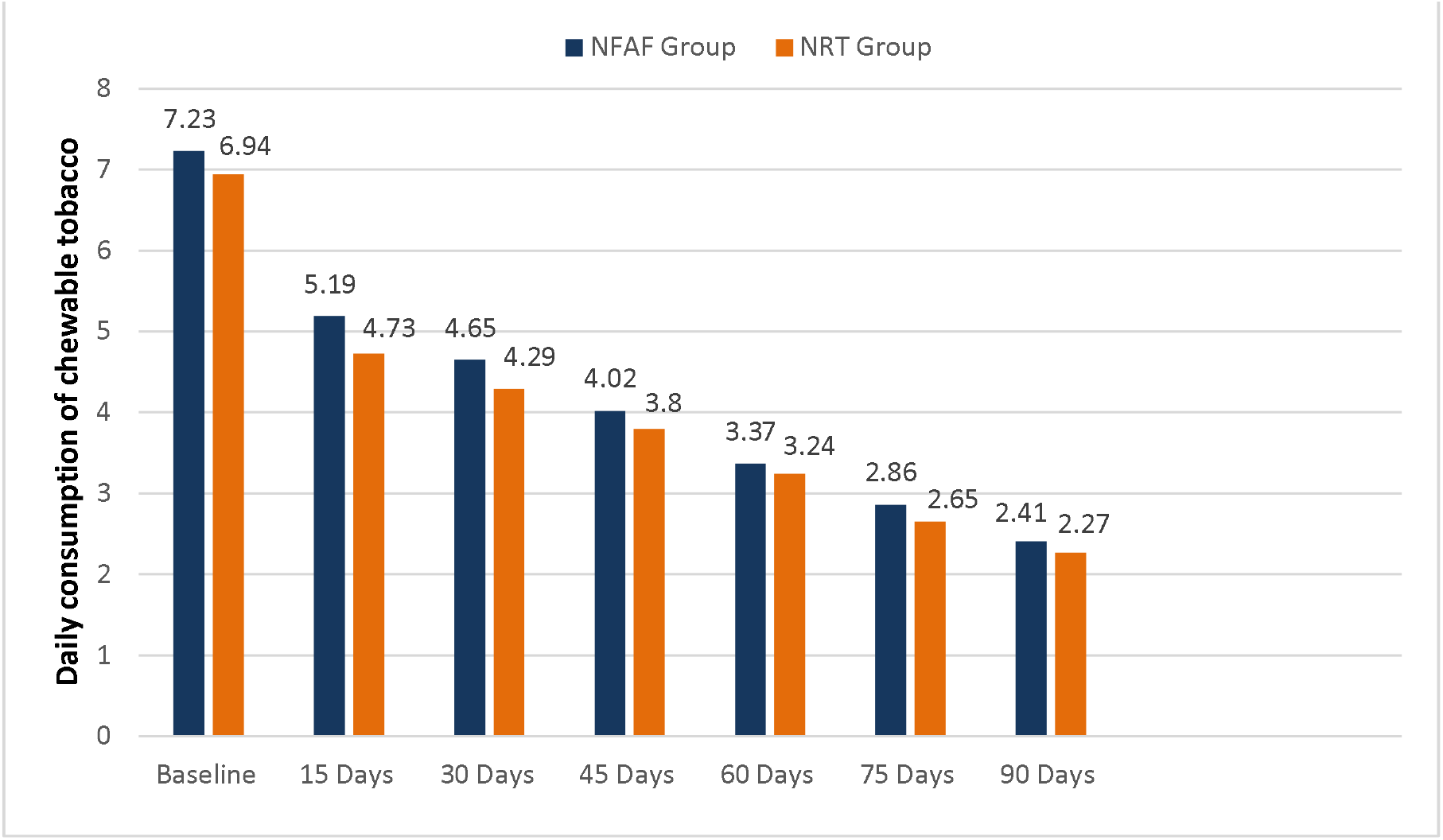
Frequency of tobacco consumption.

### Frequency with respect to no. of subjects

At the end of day 90, complete cessation of tobacco consumption was achieved by 21.56% of participants in the NFAF group, compared to 17.77% in the NRT group. Additionally, 37.35% of NFAF users and 37.77% of NRT users showed an 80–100% reduction in tobacco chewing. Across all levels of reduction, NFAF demonstrated statistically comparable efficacy to NRT.

**Table 2:**
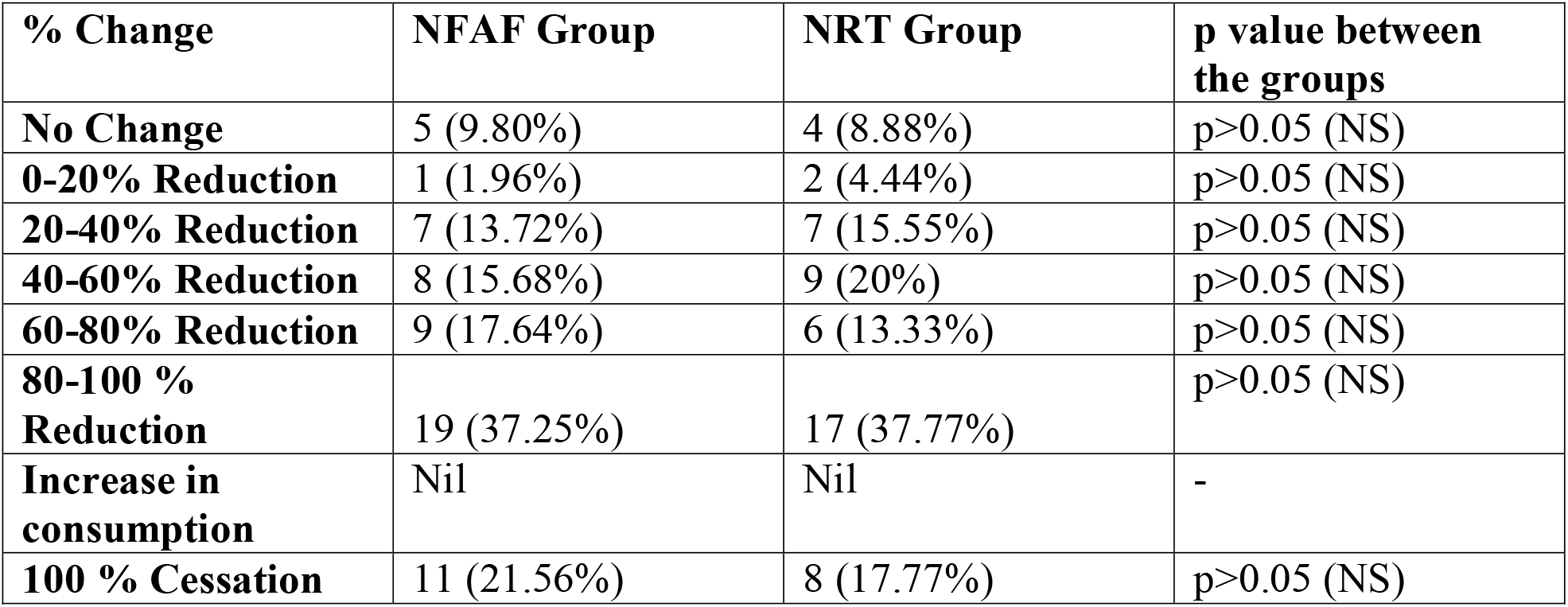
Frequency with respect to number of subjects.

### Duration of tobacco consumption per session

NFAF demonstrated a significant reduction in the average daily duration of tobacco chewing over the course of the study, with values decreasing from 14.08□±□8.71 minutes at baseline to 7.41□±□6.82 minutes at day 90 (*p*□<□0.05). This trend was comparable to the NRT group, which showed a reduction from 14.63□±□9.18 to 7.59□±□7.31 minutes. At all follow-up points, the reductions from baseline within both groups were statistically significant and comparable.

**Table 3:** Duration of tobacco consumption.

| Time point | NFAF group | NRT Group | p value between the groups |
| --- | --- | --- | --- |
| Baseline | 14.08 ± 8.71 | 14.63 ± 9.18 | p>0.05 (NS) |
| 15 Days | 14.37 ± 9.86 | 13.42 ± 9.93 | p>0.05 (NS) |
| 30 Days | 13.47 ± 10.07 | 12.70 ± 9.56 | p>0.05 (NS) |
| 45 Days | 11.55 ± 6.77* | 11.42 ± 8.97* | p>0.05 (NS) |
| 60 Days | 10.45 ± 7.13* | 10.91 ± 9.10 * | p>0.05 (NS) |
| 75 Days | 8.86 ± 7.41* | 10.14 ± 8.39* | p>0.05 (NS) |
| 90 Days | 7.41 ± 6.82* | 7.59 ± 7.31* | p>0.05 (NS) |
| *p<0.05 (Significant) from baseline to follow up visits |  |  |  |

### Mouth opening capacity

Mouth opening capacity showed a comparable, slight and non-significant increase from baseline to the end of the study in both groups.

### Quality of life

NFAF group demonstrated significant improvement in overall quality of life from baseline to study end, which was comparable to the NRT group. No significant differences between the two groups were observed in the physical and environmental health domain scores. In the psychological domain, the NFAF group showed a greater improvement, with scores increasing from 19.12□±□2.32 at baseline to 20.78□±□2.14 at day 90 (*p*□<□0.05), compared to the NRT group, which improved from 19.00□±□2.39 to 19.88□±□2.88. Both groups showed significant improvement in social health, with no significant difference observed between them at any follow-up.

**Table 4:** Quality of life.

|  | **NFAF group** |  |  |  | **NRT Group** |  |  |  |
| --- | --- | --- | --- | --- | --- | --- | --- | --- |
|  | **Baseline** | **30 Days** | **60 Days** | **90 Days** | **Baseline** | **30 Days** | **60 Days** | **90 Days** |
| **Overall quality of life** | 3.49<br>$\pm 0.67$ | 3.43<br>$\pm 0.78$ | 3.76<br>$\pm 0.55$ | 3.82<br>$\pm 0.53^*$ | 3.56<br>$\pm 0.77$ | 3.49<br>$\pm 0.77$ | 3.86<br>$\pm 0.52$ | 3.80<br>$\pm 0.40^*$ |
| **Overall satisfaction with health** | 3.27<br>$\pm 0.90$ | 3.35<br>$\pm 0.87$ | 3.73<br>$\pm 0.75$ | 3.67<br>$\pm 0.77^*$ | 3.47<br>$\pm 0.80$ | 3.53<br>$\pm 0.85$ | 3.88<br>$\pm 0.50$ | 3.90<br>$\pm 0.66^*$ |
| **Physical health** | 22.27<br>$\pm 2.19$ | 21.92<br>$\pm 2.41$ | 22.84<br>$\pm 1.98$ | 22.59<br>$\pm 2.70$ | 22.33<br>$\pm 2.24$ | 22.23<br>$\pm 2.30$ | 22.84<br>$\pm 2.21$ | 23.12<br>$\pm 2.45$ |
| **Psychological health** | 19.12<br>$\pm 2.32$ | 19.04<br>$\pm 2.30$ | 20.06<br>$\pm 1.98$ | 20.78<br>$\pm 2.14^*$ | 19.00<br>$\pm 2.39$ | 18.56<br>$\pm 2.92$ | 19.77<br>$\pm 2.64$ | 19.88<br>$\pm 2.88$ |
| **Social relationships** | 10.22<br>$\pm 1.94$ | 10.35<br>$\pm 1.98$ | 10.98<br>$\pm 1.54$ | 11.27<br>$\pm 1.59$ | 10.42<br>$\pm 1.62$ | 10.37<br>$\pm 1.89$ | 11.23<br>$\pm 1.60$ | 11.34<br>$\pm 1.49$ |
| **Environment health** | 25.49<br>$\pm 4.04$ | 26.06<br>$\pm 4.78$ | 27.88<br>$\pm 3.50$ | 28.59<br>$\pm 3.79^*$ | 26.33<br>$\pm 4.06$ | 26.70<br>$\pm 4.46$ | 28.63<br>$\pm 2.78$ | 28.49<br>$\pm 2.92^*$ |
|  | $p < 0.05$ (Significant) on psychological health domain at day 90 between the groups<br>$^*p < 0.05$ (Significant) within the group from baseline to follow up visits |  |  |  |  |  |  |  |

### Perceived stress and Sleep disturbances

NFAF group demonstrated significant reduction in stress levels as measured on PSS from baseline to the end of the study, comparable to the NRT group.

**Table 5:** Perceived Stress.

|  | Baseline | 30 Days | 60 Days | 90 Days |
| --- | --- | --- | --- | --- |
| **NFAF Group** | 17.49 ±4.68 | 17.41 ±4.95 | 15.67 ±4.42* | 15.67 ±4.09* |
| **NRT Group** | 16.84 ±5.42 | 16.81 ±4.96 | 15.63 ±4.65* | 15.24 ±4.22* |
| **p value between the groups** | p>0.05 (NS) | p>0.05 (NS) | p>0.05 (NS) | p>0.05 (NS) |

Sleep disturbances evaluated using ISI exhibited non-significant changes in the sleep schedule throughout the study period for both groups.

**Table 6:** Sleep disturbances.

|  | Baseline | 30 Days | 60 Days | 90 Days |
| --- | --- | --- | --- | --- |
| **NFAF Group** | 6.53 ±4.63 | 6.43 ±4.65 | 6.80 ±4.61 | 6.84 ±4.97 |
| **NRT Group** | 6.30 ±4.60 | 6.23 ±4.68 | 6.91 ±5.11 | 6.93 ±5.52 |
| **p value between the groups** | p>0.05 (NS) | p>0.05 (NS) | p>0.05 (NS) | p>0.05 (NS) |

### Physiological parameters

NFAF group exhibited significant improvement in energy, strength, and stamina from baseline to 30 days continuing through the study period, which was comparable to the NRT group. For appetite, digestion, and bowel habits, both groups did not demonstrate any significant changes.

**Table 7:** Physiological parameters.

|  | NFAF group |  |  |  | NRT Group |  |  |  |
| --- | --- | --- | --- | --- | --- | --- | --- | --- |
|  | Baseline | 30 Days | 60 Days | 90 Days | Baseline | 30 Days | 60 Days | 90 Days |
| **Energy** | 1.53±1.08 | 1.90±1.02* | 2.25±0.84* | 2.35±0.86* | 1.44±1.14 | 1.77±1.11* | 2.16±0.90* | 2.27±0.92* |
| **Strength** | 1.53±0.99 | 1.78±0.92* | 2.02±0.84* | 2.08±0.95* | 1.47±1.03 | 1.70±0.94* | 1.86±0.91* | 2.10±1.97* |
| **Stamina** | 1.35±1.21 | 1.78±1.08* | 1.88±1.09* | 1.92±1.06* | 1.26±1.26 | 1.67±1.08* | 1.79±1.01* | 2.00±0.97* |
|  | *p<0.05 (Significant) within the group from baseline to follow up visits<br>p>0.05 (non-significant) between the groups at all follow up visits |  |  |  |  |  |  |  |
| **Appetite** | 4.27±2.74 | 4.25±2.74 | 4.57±2.85 | 4.24±2.95 | 4.21±2.58 | 4.33±2.71 | 4.58±2.63 | 4.59±2.88 |
| **Digestion** | 4.37±2.61 | 4.25±2.59 | 4.24±2.89 | 4.16±2.87 | 4.93±2.61 | 4.28±2.77 | 4.28±3.04 | 4.15±2.90 |
| **Bowel Habits** | 3.92±2.62 | 3.24±2.59 | 3.31±2.77 | 3.02±2.90 | 3.84±2.96 | 3.12±2.87 | 3.53±2.75 | 3.15±2.83 |
|  | Between the groups - p>0.05 (not significant) from baseline to follow up visits for all the three parameters |  |  |  |  |  |  |  |

### Clinical symptoms related with tobacco consumption

NFAF group exhibited significant reduction in clinical symptoms such as headache, giddiness, lack of concentration, mood change, fatigue from 30 days onwards till the end of the study. This reduction was comparable to the NRT group.

**Table 8:**
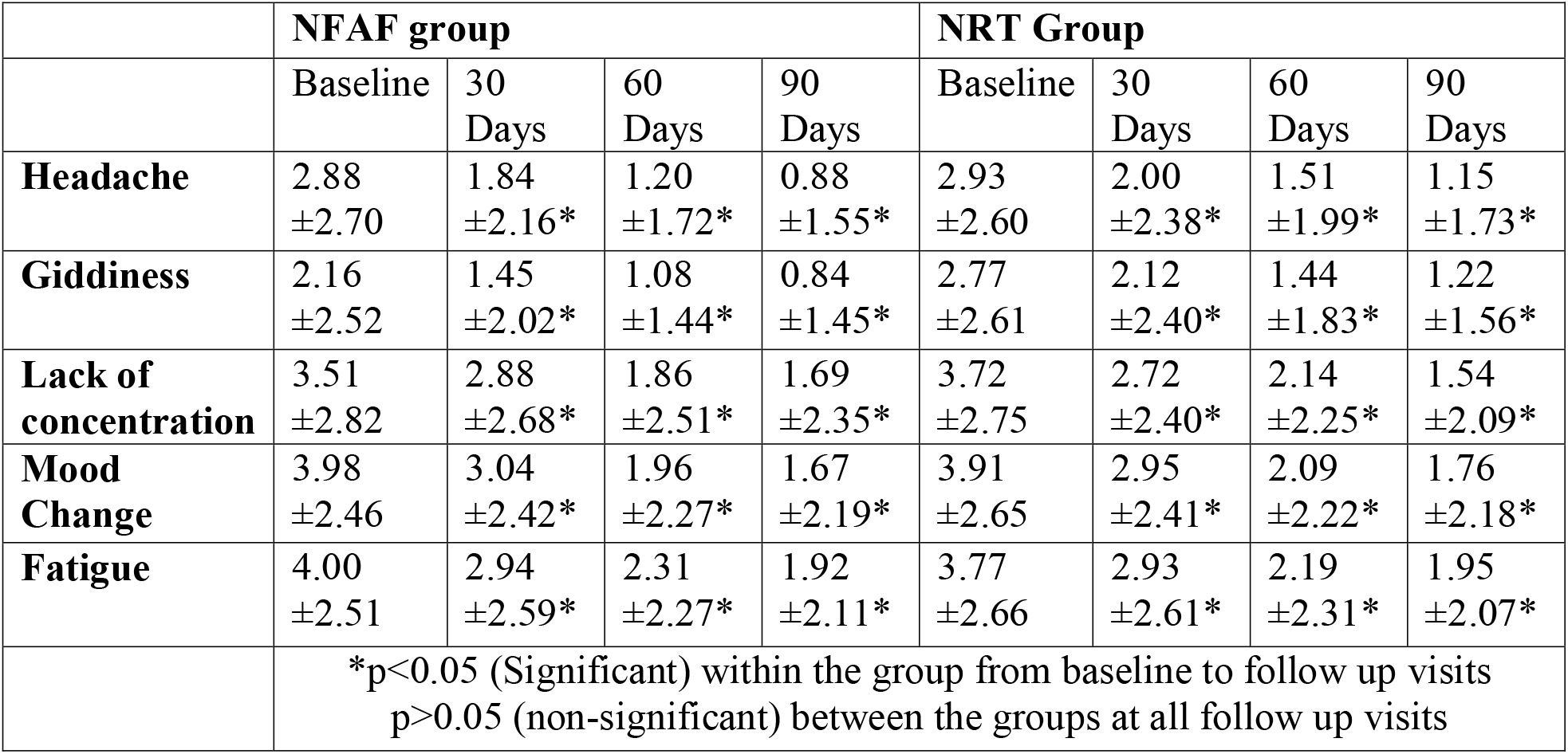
Clinical symptoms related with tobacco consumption.

|  | **NFAF group** |  |  |  | **NRT Group** |  |  |  |
| --- | --- | --- | --- | --- | --- | --- | --- | --- |
|  | Baseline | 30 Days | 60 Days | 90 Days | Baseline | 30 Days | 60 Days | 90 Days |
| **Headache** | 2.88<br>±2.70 | 1.84<br>±2.16* | 1.20<br>±1.72* | 0.88<br>±1.55* | 2.93<br>±2.60 | 2.00<br>±2.38* | 1.51<br>±1.99* | 1.15<br>±1.73* |
| **Giddiness** | 2.16<br>±2.52 | 1.45<br>±2.02* | 1.08<br>±1.44* | 0.84<br>±1.45* | 2.77<br>±2.61 | 2.12<br>±2.40* | 1.44<br>±1.83* | 1.22<br>±1.56* |
| **Lack of concentration** | 3.51<br>±2.82 | 2.88<br>±2.68* | 1.86<br>±2.51* | 1.69<br>±2.35* | 3.72<br>±2.75 | 2.72<br>±2.40* | 2.14<br>±2.25* | 1.54<br>±2.09* |
| **Mood Change** | 3.98<br>±2.46 | 3.04<br>±2.42* | 1.96<br>±2.27* | 1.67<br>±2.19* | 3.91<br>±2.65 | 2.95<br>±2.41* | 2.09<br>±2.22* | 1.76<br>±2.18* |
| **Fatigue** | 4.00<br>±2.51 | 2.94<br>±2.59* | 2.31<br>±2.27* | 1.92<br>±2.11* | 3.77<br>±2.66 | 2.93<br>±2.61* | 2.19<br>±2.31* | 1.95<br>±2.07* |
|  | *p<0.05 (Significant) within the group from baseline to follow up visits<br>p>0.05 (non-significant) between the groups at all follow up visits |  |  |  |  |  |  |  |

### Nicotine dependence using Fagerström Test

Nicotine dependence in NFAF group decreased significantly and comparably to the NRT group over the study period.

**Graph 2:**
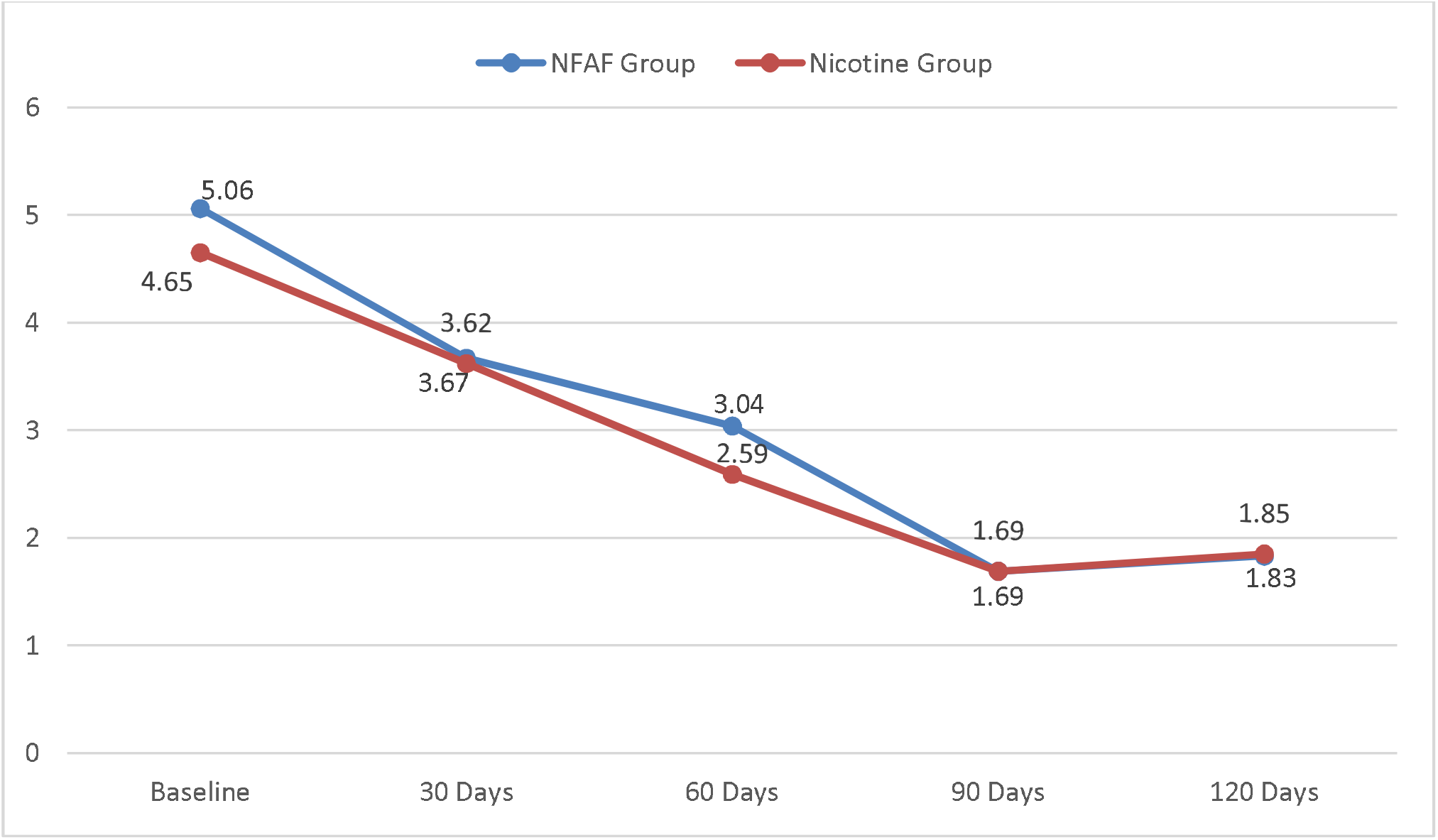
Nicotine dependence.

### Relapse

After the 90-day intervention period, participants were monitored for 30 more days to assess relapse. A higher percentage of the NFAF group (25.49%) remained tobacco-free compared to the NRT group (20.83%), however, the difference was not statistically significant.

**Table 9:** relapse.

|  | NFAF group<br>(n=51) | NRT Group (n=48) | P value between the<br>groups |
| --- | --- | --- | --- |
| **Did not**<br>**relapse** | 13 (25.49%) | 10 (20.83%) | p>0.05 (NS) |
| **Relapsed** | 38 (74.51%) | 38 (79.16%) | p>0.05 (NS) |

### Urine analysis for nicotine

At the end of the 90-day study period, 15 participants (29.42%) in the NFAF group and 12 participants (25%) in the NRT group tested negative for urinary nicotine, indicating complete cessation. While the difference was not statistically significant, the NFAF group exhibited a higher cessation rate.

### Safety related parameters (vitals, laboratory investigations, overall safety and adverse events)

Both interventions did not affect the vitals and laboratory parameters such as CBC, liver function tests, renal function tests, and blood sugar levels throughout the study period.

**Table 10:** Assessment of safety related Laboratory Investigations.

| Parameter | NFAF group |  | NRT Group |  | P value between the |
| --- | --- | --- | --- | --- | --- |
|  | Baseline | 90 Days | Baseline | 90 Days |  |
| **Hb** | 13.95±2.19 | 13.11±2.17 | 14.20±2.35 | 13.19±2.09 | p>0.05 (NS) |
| **RBC** | 4.92±0.74 | 4.69±0.75 | 5.05±0.67 | 4.81±0.75 | p>0.05 (NS) |
| **WBC** | 7282.35<br>±1773.55 | 6236.36<br>±1896.84 | 7583.72<br>±1996.66 | 6620.26<br>±1793.28 | p>0.05 (NS) |
| **Platelets** | 259019.61<br>±69043.90 | 261477.27<br>±78517.51 | 258767.44<br>±71759.13 | 266871.79<br>±81403.86 | p>0.05 (NS) |
| **ESR** | 8.32±13.38 | 8.05±11.30 | 10.58±19.75 | 8.82±14.02 | p>0.05 (NS) |
| **Total Bilirubin** | 0.83±0.39 | 0.94±0.54 | 0.80±0.42 | 0.81±0.43 | p>0.05 (NS) |
| **AST** | 59.25±28.39 | 49.59±20.99 | 57.44±24.57 | 46.15±18.85 | p>0.05 (NS) |
| **ALT** | 66.98±40.33 | 45.91±25.58 | 58.30±30.88 | 41.85±26.07 | p>0.05 (NS) |
| **Alk. Phosphatase** | 69.98±21.86 | 63.95±23.02 | 73.65±28.11 | 66.44±23.35 | p>0.05 (NS) |
| **Sr. Proteins** | 7.71±0.52 | 7.91±0.57 | 7.80±0.53 | 7.83±0.62 | p>0.05 (NS) |
| **BUN** | 9.73±2.76 | 9.95±3.25 | 10.40±2.56 | 9.45±2.61 | p>0.05 (NS) |
| **Sr. Creatinine** | 0.78±0.19 | 0.80±0.21 | 0.85±0.21 | 0.81±0.20 | p>0.05 (NS) |
| **Blood Sugar (Random)** | 86.67<br>±15.83 | 100.02<br>±22.64 | 91.72<br>±16.13 | 105.95<br>±25.13 | p>0.05 (NS) |

NFAF granules were generally rated as having good or excellent overall safety, both by the investigators and the subjects themselves. A total of 14 and 15 adverse events were recorded in NFAF and NRT groups respectively, however they were observed to be unrelated to the study product or procedure and were resolved without requiring stoppage of study medication or any sequelae. Adverse events recorded included burning micturition, chest burning, body ache, mouth ulcer, diarrhea, cold with fever, hiccups, cough and cold, body ache with fever, cough, uneasiness, headache, headache with giddiness, and giddiness.

## Discussion

SLT use surpasses all other forms of tobacco consumption in certain regions of the world. A wide variety of these products are used globally, each differing significantly in preparation, method of use, and toxicity. However, substantial evidence highlights the harmful effects of SLT products, particularly in the Asia and Middle East region, where their use is strongly associated with increased mortality — especially from cancer, coronary heart disease (CHD), and overall causes. While the link to respiratory mortality is less pronounced, use of these products is clearly linked to higher CVD morbidity, with strong associations to ischemic heart disease and stroke. Overall, the health risks associated with SLT use remain significant and concerning.^15^

In this open-label, randomized, multicentric clinical trial, the proposed NFAF demonstrated significant improvements in tobacco consumption frequency, duration, and nicotine dependence over 90 days, which were comparable with the conventional NRT. Both groups were comparable across demographic or baseline characteristics. NFAF demonstrated improvement comparable to NRT in levels of perceived stress (PSS), energy, strength, stamina, and quality of life. Reduction of clinical symptoms associated with SLT consumption was also comparable in both groups. Tobacco chewing cessation rates achieved by NFAF were slightly higher than NRT, although the difference was not statistically significant.

Adverse events were minor, resolved without sequelae, and were not attributed to either intervention. Safety assessments rated both products favourably, with most participants and investigators grading safety as good to excellent. Overall, NFAF granules were well-tolerated and achieved efficacy outcomes comparable to standard NRT, supporting their potential as a safe and effective herbal option for tobacco cessation.

Proposed NFAF incorporates a synergistic blend of ingredients aimed at addressing the multifaceted challenges of cessation of chewing tobacco. The combination provides dopaminergic support through *Mucuna pruriens* to mitigate mood disturbances and reduce cravings. Adaptogenic and anxiolytic agents such as *Withania somnifera, Ocimum sanctum*, and green tea help alleviate stress, anxiety, and sleep disturbances associated with withdrawal.

Ingredients like *Lavang*, guava, and green tea contribute to oral health through their anti-inflammatory and antimicrobial properties, aiding in the healing of tobacco-related oral damage. *Akalkara* and *Supari* offer strong sensory and stimulant effects that may act as substitutes for the oral fixation and psychoactive stimulation of tobacco, while *Amaltas* supports gastrointestinal comfort during the withdrawal phase. Together, the formulation addresses neurochemical imbalance, stress, oral health, and behavioural habits, offering a comprehensive approach to deaddiction. ^16–24^

## Conclusion

The findings of this study indicate that the proposed Nicotine free Ayurvedic formulation is effective in reducing oral chewable tobacco addiction and facilitating complete cessation in a subset of participants with efficacy comparable to NRT. In addition to reducing tobacco dependence, this formulation also caused significant improvements in quality of life, energy, stamina, and stress levels. The formulation was well tolerated, with no adverse effects reported, and laboratory parameters remained within normal limits throughout the study period. These results suggest that the proposed Nicotine free Ayurvedic formulation may serve as a safe and effective herbal alternative to conventional NRT for tobacco cessation.

## Data Availability

All data produced in the present study are available upon reasonable request to the authors

